# Elevated serum uric acid is a facilitating mechanism for insulin resistance mediated accumulation of visceral adipose tissue

**DOI:** 10.1101/2020.09.20.20198499

**Authors:** Luisa Fernández-Chirino, Neftali Eduardo Antonio-Villa, Carlos Alberto Fermín-Martínez, Alejandro Márquez-Salinas, Enrique C. Guerra, Arsenio Vargas-Vázquez, Paloma Almeda-Valdés, Donají Gómez-Velasco, Tania Leticia Viveros-Ruiz, Rosalba Rojas, Carlos A. Aguilar Salinas, Omar Yaxmehen Bello-Chavolla

## Abstract

**OBJECTIVE:** Serum uric acid (SUA) has been associated to cardiometabolic conditions such as insulin resistance (IR) and visceral adipose tissue (VAT) accumulation. Here, we aimed to clarify a unifying mechanism linking elevated SUA to IR and VAT.

**METHODS:** We conducted analyses in 226 subjects from the UIEM cohort with both euglycemic hyperinsulinemic clamp (EHC) and dual X-ray absorptiometry (DXA) measurements for IR and VAT accumulation, and explored the role of SUA and adiponectin by developing a network of causal mediation analyses to assess their impact on IR and VAT. These models were then translated to two population-based cohorts comprising 6,337 subjects from NHANES 2003-2004 and 2011-2012 cycles in the US and ENSANUT Medio Camino 2016 in Mexico, using HOMA2IR and adipoIR as indicators of peripheral and adipose tissue IR, and METS-VF as a surrogate for VAT accumulation.

**Results:** SUA has a mediating role inside a bidirectional relationship between IR and visceral obesity, which was similar using either gold standard measurements or surrogate measures for IR and VAT. Furthermore, adiponectin acts as a linking mediator between elevated SUA and both peripheral IR and VAT accumulation. The proportion of the mechanism for IR-mediated (in either peripheral or adipose tissue) VAT accumulation was greater, compared to VAT-mediated IR accumulation (10.53%[9.23%-12.00%] to 5.44%[3.78%-7.00%]). Normal-range SUA levels can be used to rule-out underlying cardio-metabolic abnormalities in both men and women.

**CONCLUSIONS:** Elevated SUA acts as mediator inside the bidirectional relationship between IR and VAT accumulation and these observations could be applicable at a phenotype scale.

## INTRODUCTION

Uric acid is a heterocyclic puric compound and the final product of purine oxidative metabolism^1,2^. Serum uric acid (SUA) levels are dependent on age and sex; they are also associated with chronic kidney disease and its progression, probably due to its decreased renal excretion^2^. Impairments in urate metabolism are related to several cardio-metabolic conditions including cardiovascular disease (CVD), nephrolithiasis, gout, arterial hypertension, dyslipidemias, and loss of plasmatic antioxidant capacity^3–5^.

Two conditions which have been linked to SUA-related metabolic impairments include insulin resistance (IR) and visceral adipose tissue (VAT) accumulation. Independently, IR and VAT accumulation have a bidirectional relationship^6^: IR can be caused by metabolic dysregulation due to excessive VAT accumulation and at the same time, IR can cause this excessive VAT accumulation^6,7^. Both phenomena have been studied as interlinked conditions to SUA metabolism through different pathophysiological pathways in in vivo models and humans mainly involving oxidative stress, electrolyte equilibrium, immunometabolic regulators and specific enzymatic deregulation^8–10^. Nevertheless, SUA has only been hypothesized to be a part of an underlying causality mechanism linking both IR and VAT in *in* vitro studies, with in vivo results yielding similar results^9^. Additionally, VAT has significant metabolic and endocrine activity and many of its deleterious effects on cardio-metabolic health have been postulated to be promoted by specific adipokine secretory profiles, particularly adiponectin and leptin^11^.

Mechanistic, population-based studies focused in determining the nature of the relationship between SUA and both IR and VAT accumulation are currently scarce. Identifying a mediating role of SUA in both IR and VAT accumulation will ultimately clarify the role of elevated SUA in cardio-metabolic diseases and evaluate its utility as a complementary measure to assess metabolic health. Here, we attempt to clarify a mediating pathophysiological pathway relating VAT accumulation and IR, where SUA acts as a link between both phenomena using national-based surveys in two countries.

## METHODS

### UIEM Cohort

We included Mexican subjects (n=226) from the UIEM cohort, which comprises individuals from the SIGMA study (n=129) together with a subset of non-diabetic individuals (n=97) with either obesity (BMI≤40kg/m^2^) or normal weight (BMI*<*25kg/m^2^)^12^. In this cohort, insulin sensitivity was assessed using euglycemic hyperinsulinemic clamp (EHC), VAT volume using Dual X-Ray Absorptiometry (VAT-DXA), and plasma adiponectin was measured using ELISA assays (Merck Millipore). The Human Research Ethics Committee of the Instituto Nacional de Ciencias Médicas y Nutrición Salvador Zubirán approved the study, and all participants in the cohort provided written consent after the full explanation of the purpose and nature of all procedures used.

### Population-based Cohorts

We analyzed, both separately and jointly, an ethnically diverse cohort comprised of Mexican (ENSANUT Medio Camino 2016) and American (NHANES 2003-2004 and 2011-2012 cycles) subjects. The goal of both cohorts was to assess the health and nutritional status in each country based on a comprehensive stratified sample of subjects from each population. Further sampling and stratification methodology and methods for both ENSANUT and NHANES cohorts are published elsewhere^13–15^. A detailed flowchart of sample selection is depicted in **Supplemental Material**. Briefly, out of both NHANES cycles, we selected subjects with insulin and palmitate measurements for the cohort analysis. For ENSANUT we selected subjects with complete uric acid measurements. The Human Research Ethics Committee of the Instituto Nacional de Ciencias Médicas y Nutrición Salvador Zubirán approved the study.

### Laboratory and anthropometric measurements

For the NHANES, anthropometric and biochemical measurement methods, are specified for both 2003-2004 and 2011-2012 cycles elsewhere; all these subjects had fasting plasmatic free fatty acid quantifications. For ENSANUT and UIEM cohorts, anthropometric measurements, such as height, weight, and waist circumference were measured by trained professionals to one significant digit; biochemical measurements were obtained after a 10-12h fasting period and assessed in a centralized location. BMI was categorized as underweight: *<*18.5 kg/m^2^, normal-weight: 18.5-25.0 110 kg/m^2^, overweight: 25.1 - 29.9 kg/m^2^, obesity: 30.0 - 39.9 kg/m^2^, extreme obesity: *>*40 kg/m^2^. Estimated glomerular filtration rate (eGFR) was calculated using the CKD-EPI equation. We also analyzed data on drug use information including use of antihypertensives, antihyperuricemics, aspirin, and lipid-lowering drugs.

### Definitions of metabolic conditions

For the UIEM cohort, IR was defined as M-values [mg/min/kg] from EHC ≤4.7^16^. For NHANES and ENSANUT, IR was estimated using the HOMA2IR, obtained along with HOMA2%B and HOMA2%S calculator using glucose and insulin values obtained in a fasting state of 8-to-12 hrs, and calculated using the Oxford Centre Diabetes Trial Unit calculator^17^. Peripheral IR was defined as values *>*80th percentile for HOMA2IR, which was *>*2.02 for NHANES and *>*2.15 for ENSANUT. To explore the role of adipose tissue IR, we calculated the Adipo_palmitate_IR index as: AdipoIR=Palmitate [*µmol/L*] × Fasting insulin [*pmol/L*]) × 4. Palmitate accounts for approximately one-fourth of total plasmatic free fatty acids, therefore, Adipo_palmitate_IR was selected as a measure of adipose tissue insulin resistance to ensure better reproducibility. Adipose tissue IR was defined with sex-specific cut-off points for adipoIR >75^th^ percentile as previously described^35^, which were >3.05 for males and *>*3.06 for females^18^. In the UIEM cohort, visceral obesity was defined as VAT-DXA *>* 1000g. For ENSANUT and NHANES, we estimated VAT using the Metabolic Score for Visceral Fat (METS-VF)^19^, calculated as:

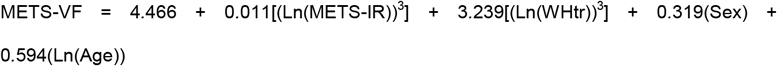

Visceral obesity was defined as METS-VF>7.18.

### Statistical analysis

Categorical variables are presented as frequencies and continuous variables as mean (±standard deviation) or median (interquartile range), wherever appropriate. Differences between groups were tested using Chi-Squared tests for categorical variables and Mann-Whitney U or t-tests for continuous variables, wherever appropriate. Kruskal-Wallis tests with Dunn tests for pairwise comparisons were used and corrected for multiple comparisons. To evaluate whether significant discrepancies were observed between ENSANUT and NHANES cohorts, we used Principal Component Analysis (PCA). Variable transformations were applied to approximate a normality in all analyzed variables using logarithmic, ordered quantile (ORQ) normalization or square root transformations wherever appropriate. All statistical analyses and data management were done with R version 4.0.0. A p value <0.05 was considered as the statistical significance threshold.

### Missing data

Missing data was dealt with using multiple imputation with chained equations under the assumption of data missing completely at random with the *mice* R package, developing 5 multiply imputed datasets for a maximum of five iterations Imputed datasets were combined using Rubin’s rules. Details of imputed variables are shown in **Supplemental Material**.

### Association of SUA with IR and VAT

To establish that SUA has a correlation with IR and VAT, we used Spearman Correlation tests for each cohort separately. Regarding surrogate VAT indicators, we assessed both METS-VF and the Waist-to-Height Ratio (WHtR) to demonstrate similarities in correlation between all variables of interest and both measures as a sensitivity analysis. We also assessed these associations using multiple linear regression models by using SUA, IR and VAT either dependent or predictor variables to evaluate all possible directions for these associations independently in ENSANUT and NHANES. For the analysis of joint cohorts, we fitted mixed effects linear regression models, considering corresponding cohort of origin as a random intercept to account for potential clustering. Models were adjusted by age, sex, ethnicity, and serum creatinine or eGFR and were selected using minimization of the Bayesian Information Criteria (BIC). Assumptions were verified by analyzing standardized residuals.

### SUA mediation in IR and VAT accumulation

We hypothesized that SUA would act as a mediator of the bidirectional relationship between peripheral/adipose tissue IR and VAT accumulation. To assess this hypothesis, we conducted mediation analyses using the *mediation* R package to estimate average direct effects (ADE), average causal mediation effects (ACME) and the proportion of mediation. Inference on model estimates were performed using bootstrapping (B=1,000) to estimate 95% confidence intervals with the percentile method in adjusted analyses.

### Joint mediator between SUA and adiponectin

For mediation analyses in the UIEM cohort, we included adiponectin both as a separate additional mediator and joint together with SUA to assess its combined role within the causal mediation framework. This joint mediator between adiponectin and SUA was calculated as:

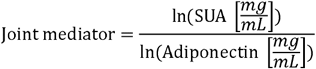

### Diagnostic performance of SUA

To evaluate SUA as an indicator of peripheral and adipose tissue IR and VAT accumulation, we used the area under the receiver operating characteristic (AUROC) curves with the *pROC* R package, in NHANES and ENSANUT separately. We also estimated cut-off points to identify these phenotypes using the *OptimalCutpoints* R package with the Youden Index to estimate sensitivity, specificity, positive and negative predictive values, and likelihood ratios.

## RESULTS

### Study Populations

We conducted all analyses in the UIEM cohort and further extrapolated them onto the population-based analysis comprising 6,316 subjects. Subjects with elevated SUA had higher levels of glucose, insulin, triglycerides, BMI, WHtR, creatinine, HOMA2%B, METS-VF and HOMA2IR levels, but lower levels of eGFR, high-density lipoprotein cholesterol (cHDL), and HOMA2%S. Similarly, cases with elevated SUA had lower levels of adiponectin and M-FFM and higher levels of leptin and VAT mass assessed by DXA. When comparing cases with normal and elevated SUA in the joint cohort, we observed similar trends to those observed in the UIEM cohort (**Table 1**). For NHANES, there were no differences in sex, antihyperuricemic, aspirin, and antihyperlipidemic use. ENSANUT showed differences in all variables between elevated and normal SUA groups except for age and cases with diabetes. The biplot obtained to evaluate possible discrepancies in measurements due to cohort of origin as a result of the PCA analysis is presented in **Supplementary Material**; this confirmed that ENSANUT and NHANES could be jointly evaluated using a mixed-effects framework.

**Table 1.** General characteristics of studied cohort (NHANES-ENSANUT). Patients with elevated SUA are those with a serum concentration greater than 288.48 umol/L for females and 356.88 umol/L for males. Values are presented as means (standard deviation), or as frequencies, where appropriate. *Abbreviations:* BMI: Body Mass Index. cHDL: Cholesterol-high density lipoprotein. HOMA2IR: Homeostatic Model for Insulin Resistance. HOMA2%S: Homeostatic Model for Insulin Resistance for pancreatic β cell sensitivity. HOMA2%B: Homeostatic Model for Insulin Resistance for functionality of pancreatic β cells. METS-VF: Metabolic Score for Visceral Fat. WHtR: Waist-height ratio; EHC: Euglycemic Hyperinsulinaemic Clamp; DXA: Dual X-ray Absorciometry

|  | UIEM cohort | Normal SUA | Elevated SUA | *p*-values | NHANES-ENSANUT cohort | Normal SUA | Elevated SUA | *p*-values |
| --- | --- | --- | --- | --- | --- | --- | --- | --- |
| n | 226 | 132 | 94 |  | 6337 | 3210 | 3106 |  |
| Age [years] | 39.84<br>($\pm 14.70$) | 36.60 ($\pm 14.16$) | 43.01<br>($\pm 14.49$) | 0.001 | 46.00<br>(30.00) | 44.00<br>(29.00) | 49.00<br>(30.00) | <0.001 |
| Female sex | 53<br>(23.45%) | 27 (20.45%) | 26<br>(27.66%) | 0.001 | 2888<br>(45.73%) | 1388<br>(43.24%) | 1500<br>(48.29%) | 0.037 |
| Glucose [mmol/L] | 5.48<br>($\pm 1.22$) | 5.32 ($\pm 1.06$) | 5.64<br>($\pm 1.33$) | 0.045 | 5.38 (0.94) | 5.28<br>(0.85) | 5.49<br>(0.95) | <0.001 |
| Insulin [pmol/L] | 58.34<br>(59.73) | 43.41 (31.08) | 79.17<br>(72.23) | <0.001 | 56.34<br>(58.33) | 47.23<br>(46.09) | 69.18<br>(67.01) | <0.001 |
| Uric acid [$\mu\text{mol/L}$] | 316.54<br>($\pm 72.89$) | 262.59($\pm 45.47$) | 366.33<br>($\pm 56.04$) | <0.001 | 315.24<br>(113.01) | 261.71<br>(71.38) | 374.72<br>(89.22) | <0.001 |
| TGL [mmol/L] | 1.36<br>(0.84) | 1.21 (0.73) | 1.51<br>(0.84) | <0.001 | 1.40 (1.15) | 1.23<br>(1.01) | 1.59<br>(1.22) | <0.001 |
| cHDL [mmol/L] | 1.17<br>($\pm 0.31$) | 1.27 ($\pm 0.33$) | 1.08<br>($\pm 0.25$) | <0.001 | 1.22 (0.52) | 1.27<br>(0.54) | 1.14<br>(0.44) | <0.001 |
| Total cholesterol [mmol/L] | 4.60<br>(±0.87) | 4.49 (±0.83) | 4.71<br>(±0.89) | **0.059** | 4.89 (1.37) | 4.84<br>(1.37) | 4.97<br>(1.37) | <0.001 |
| BMI [kg/m<sup>2</sup>] | 31.29<br>(±10.50) | 27.48 (±7.98) | 34.87<br>(±11.33) | <0.001 | 27.66<br>(7.50) | 26.09<br>(6.87) | 29.19<br>(7.52) | <0.001 |
| WHtR | 0.72<br>(±0.17) | 0.68 (±0.15) | 0.75<br>(±0.19) | 0.002 | 0.59 (0.12) | 0.56<br>(0.12) | 0.61<br>(0.12) | <0.001 |
| Creatinine [mmol/L] | 60.90<br>(±72.89) | 262.59<br>(±45.47) | 366.33<br>(±56.04) | **0.533** | 70.72<br>(24.75) | 66.30<br>(23.86) | 74.26<br>(1.37) | <0.001 |
| eGFR [mL/min/1.73m<sup>2</sup>] | 110.21<br>(±17.15) | 113.54<br>(±15.32) | 107.14<br>(±18.21) | 0.005 | 101.15<br>(30.77) | 105.57<br>(27.52) | 95.82<br>(33.57) | <0.001 |
| HOMA2%B | 91.40<br>(50.40) | 80.30 (42.70) | 101.30<br>(59.00) | <0.001 | 81.30<br>(55.00) | 74.60<br>(48.32) | 89.60<br>(57.50) | <0.001 |
| HOMA2%S | 88.60<br>(85.00) | 122.15 (77.23) | 65.90<br>(62.80) | <0.001 | 91.50<br>(92.30) | 109.50<br>(105.75) | 74.90<br>(75.25) | <0.001 |
| HOMA2IR | 1.13<br>(1.16) | 0.82 (0.64) | 1.52<br>(1.39) | <0.001 | 1.09 (1.15) | 0.91<br>(0.93) | 1.34<br>(1.30) | <0.001 |
| Insulin Resistant [HOMA2IR] | 45<br>(19.91%) | 12 (9.09%) | 33<br>(35.11%) | <0.001 | 1261<br>(19.97%) | 436<br>(13.58%) | 825<br>(26.56%) | <0.001 |
| Subjects with diabetes [n, %] | 53<br>(23.45%) | 21 (15.91%) | 32<br>(34.04%) | 0.001 | 342<br>(5.41%) | 124<br>(3.86%) | 218<br>(7.02%) | <0.001 |
| METS-VF | 7.12<br>(±0.84) | 6.78 (±0.98) | 7.43<br>(±0.52) | <0.001 | 6.97 (0.89) | 6.77<br>(1.02) | 7.14<br>(0.69) | <0.001 |
| Visceral obesity [METS-VF] | 141<br>(62.39%) | 53 (40.15%) | 88<br>(93.62%) | <0.001 | 2173<br>(34.40%) | 797<br>(24.83%) | 1376<br>(44.30%) | <0.001 |
| Mexicans [n, %] | - | - | - | - | 1914<br>(30.30%) | 906<br>(28.22%) | 1008<br>(32.45%) | 0.020 |
| Mexican-Americans [n, %] | - | - | - | - | 657<br>(10.40%) | 385<br>(11.99%) | 272<br>(8.76%) | <0.001 |
| Non-hispanic | - | - | - | - | 1968 | 979 | 989 | **0.822** |
| White [n,%] |  |  |  |  | (31.16%) | (30.50%) | (31.84%) |  |
| Non-hispanic Black [n,%] | - | - | - | - | 928<br>(14.69%) | 467<br>(14.55%) | 461<br>(14.84%) | **0.844** |
| Other ethnicity [n,%] | - | - | - | - | 849<br>(13.44%) | 473<br>(14.74%) | 376<br>(12.11%) | 0.001 |
| Adiponectin [µg/mL] | 7.71<br>(5.25) | 9.45 (5.99) | 7.07<br>(4.18) | 0.001 | - | - | - | - |
| Leptin [µg/mL] | 18.45<br>(35.63) | 16.93 (17.01) | 23.34<br>(76.65) | 0.005 | - | - | - | - |
| M-value [mg/min/kg] | 5.97<br>(4.66) | 7.16 (4.56) | 4.56<br>(4.43) | <0.001 | - | - | - | - |
| IR-EHC [M-value>4.6] | 86<br>(38.05%) | 26 (11.50%) | 60<br>(26.55%) | <0.001 | - | - | - | - |
| VAT mass [g] | 1163.12<br>(±851.29) | 817.68<br>(±734.43) | 1458.36<br>(±835.64) | <0.001 | - | - | - | - |
| Visceral obesity [DXA>1000g] | 118<br>(52.21%) | 36 (27.27%) | 82<br>(87.23%) | <0.001 | - | - | - | - |

### SUA levels in IR and visceral obesity

We observed a positive correlation between SUA and M-FFM and VAT-DXA in the UIEM cohort and with HOMA2IR, METS-VF, and the WHtR in all evaluated cohorts (**Supplementary Material**). SUA levels were higher for ascending BMI categories; notably, these trends were independent of visceral obesity and IR status. When assessing subjects according to concordance of visceral obesity and peripheral IR, we observed a similar trend for increased SUA levels for subjects with only IR or visceral obesity, or with both (**Figure 1**).

**Figure 1.**
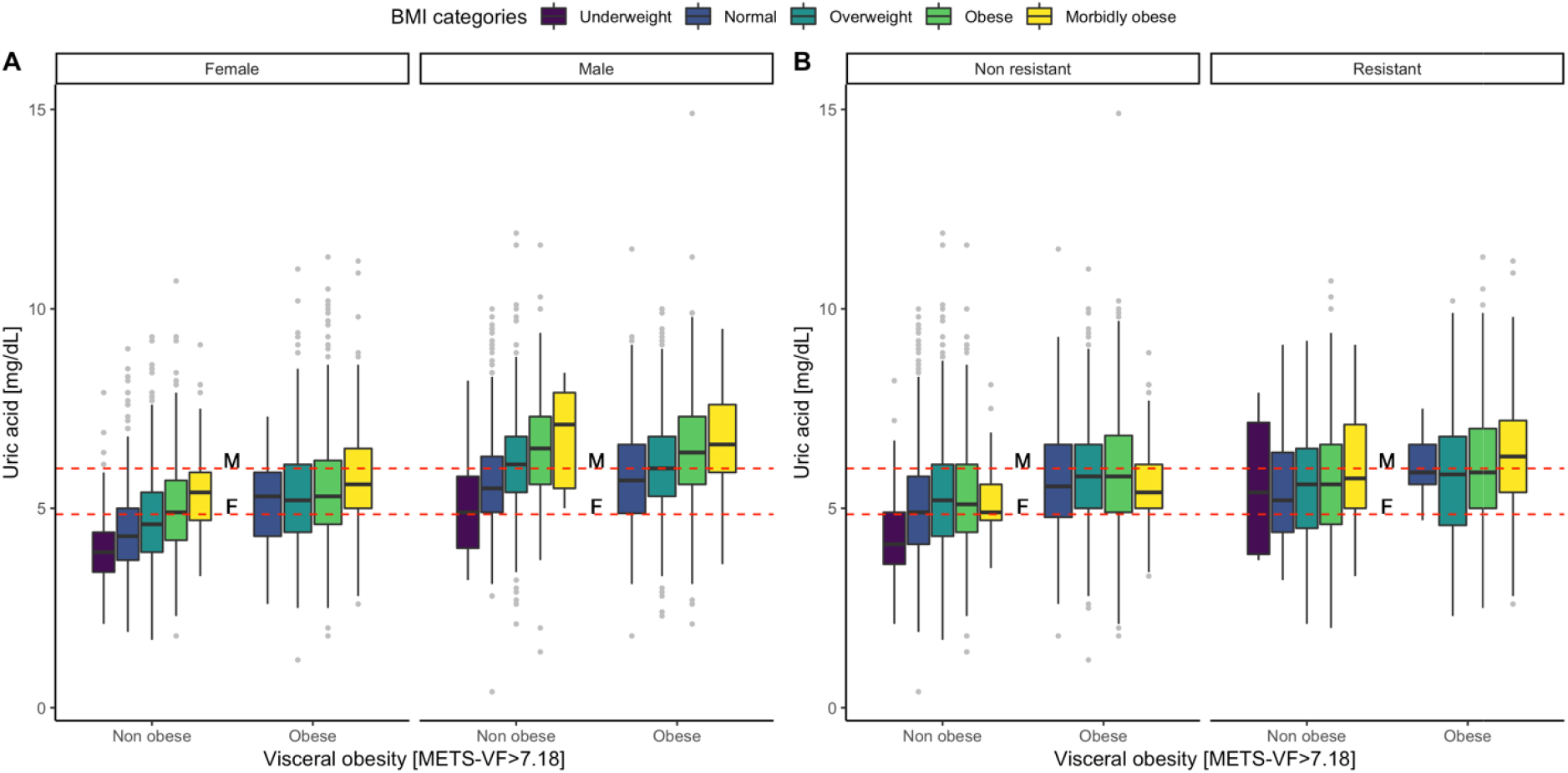
Box plot graph comparing serum uric acid values comparing cases with and without insulin resistance defined using HOMA2-IR and visceral obesity defined using METS-VF in male and female participants in the NHANES-ENSANUT joint cohort. Horizontal lines represent limit for elevated SUA in males (A, 356.88 umol/L) and in females (B, 288.48 umol/L). *Abbreviations*: HOMA2IR: Homeostatic Assessment Model for Insulin Resistance. METS-VF: Metabolic Score for Visceral Fat.

### SUA as a predictor for IR and visceral adiposity

We fitted linear regression models assessing every possible direction in the relationship between SUA, IR, and visceral fat (**Supplementary Material**). Associations between individual variables were as strong when using M-values from EHC or HOMA2IR as a peripheral IR marker or adipoIR as an adipose tissue IR marker. This suggested that elevated SUA was involved in the association between tissue-specific IR. SUA levels were also predictive of VAT-DXA in the UIEM cohort and METS-VF in the population-based cohort. Notably, the association between SUA and both IR modalities was attenuated when adjusting for VAT and vice versa for VAT when adjusted for IR modalities, indicating that SUA may act as a mediator in this relationship (**Supplementary Material**). Therefore, we formulated a bidirectional mediation model which hypothesized that elevated SUA could act as a mediator in peripheral and adipose tissue IR-mediated VAT accumulation (**Figure 2A**).

**Figure 2.**
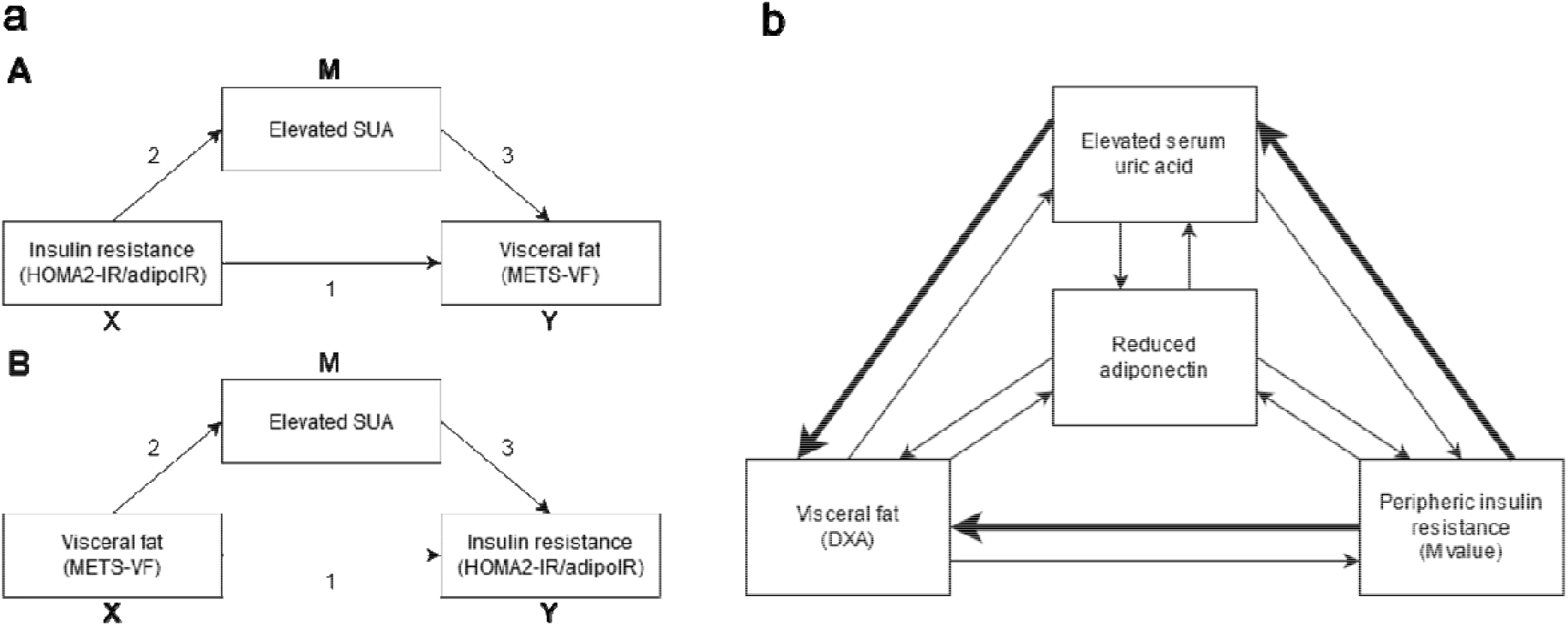
Mediation diagrams depicting general (a) and confirmatory (b) mediation analyses to explore the mediating role of serum uric acid inside the bidirectional relationship between insulin resistance and visceral adipose tissue accumulation. *Abbreviations*: adipoIR: Adipose Insulin Resistance index. DXA: Dual X-Ray absorptiometry. HOMA2IR: Homeostatic Assessment Model for Insulin Resistance. SUA: Serum uric acid.

### SUA is a mediator between VAT accumulation and peripheral or adipose tissue IR

Guided by our hypothesized mediation model, a total of four mediation pathways were investigated: 1) SUA as a mediator of peripheral IR-promoted VAT accumulation, 2) SUA as mediator or adipose tissue IR-promoted VAT accumulation, 3) SUA as a mediator of VAT-promoted peripheral IR and 4) SUA as a mediator of VAT-promoted adipose tissue IR. All four models were explored independently in NHANES and models 1) and 3) were evaluated in the UIEM cohort and confirmed in the joint cohort. First, we sought to confirm our hypothesis of the existence of a bidirectional relationship between peripheral IR and VAT accumulation in the UIEM cohort. We observed that 10.48% (95%CI 2.91-19.00%) of the effect of IR-promoted VAT accumulation was mediated by elevated SUA and that 12.91% of the effect of VAT-promoted peripheral IR (95ci 5.12-21.00%), was also likely mediated by elevated SUA levels. Because the UIEM cohort is comprised of highly selected individuals and may not represent metabolic trends of the overall population, we sought to validate these findings using population-based analyses^12^. In the joint cohort, the hypothesis which stated that peripheral IR leads to VAT accumulation showed a greater mediated proportion than the opposite direction (10.56% [95%CI 19.23.12.00%] vs. 5.44% [95%CI 3.78-7.00%], **Table 2**). However, both directions describing the mediating role of SUA in the mechanism were significant. Specifically, for NHANES, we additionally conducted all mediation models using adipoIR as a surrogate of adipose tissue IR. The observed mediated proportion was similarly stronger for adipose tissue IR-promoted VAT accumulation compared to the opposite direction (13.20% [95%CI 11.60-15.00%] vs 10.19% [95%CI 7.98-12.00%], **Table 2)**. Models conducted using logistic regressions models for phenotypes of categorized variables confirmed all observed associations, indicating that this mechanism can also be observed when the metabolic phenotypes are already established. Based on results from all models integrated at different tissue levels, we propose that elevated SUA acts as a mediator of adipose tissue/peripheral IR promoted VAT accumulation.

**Table 2.**
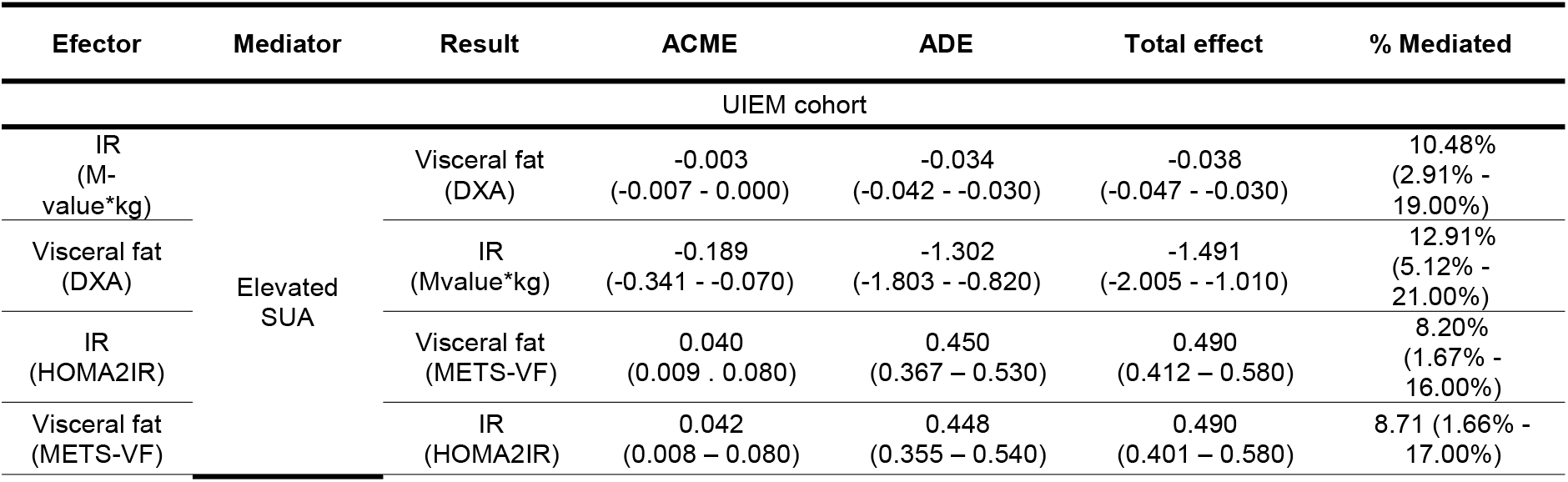
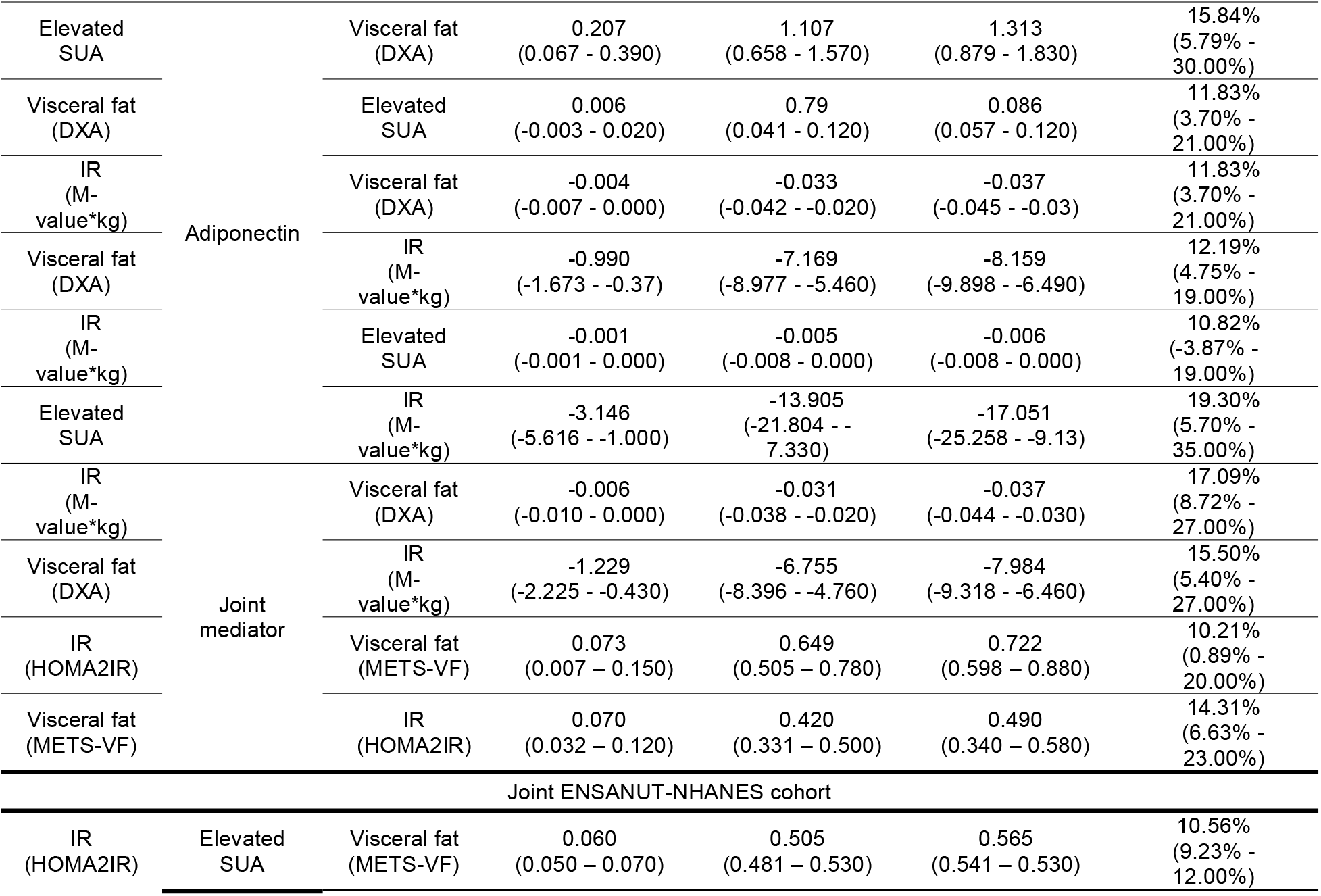
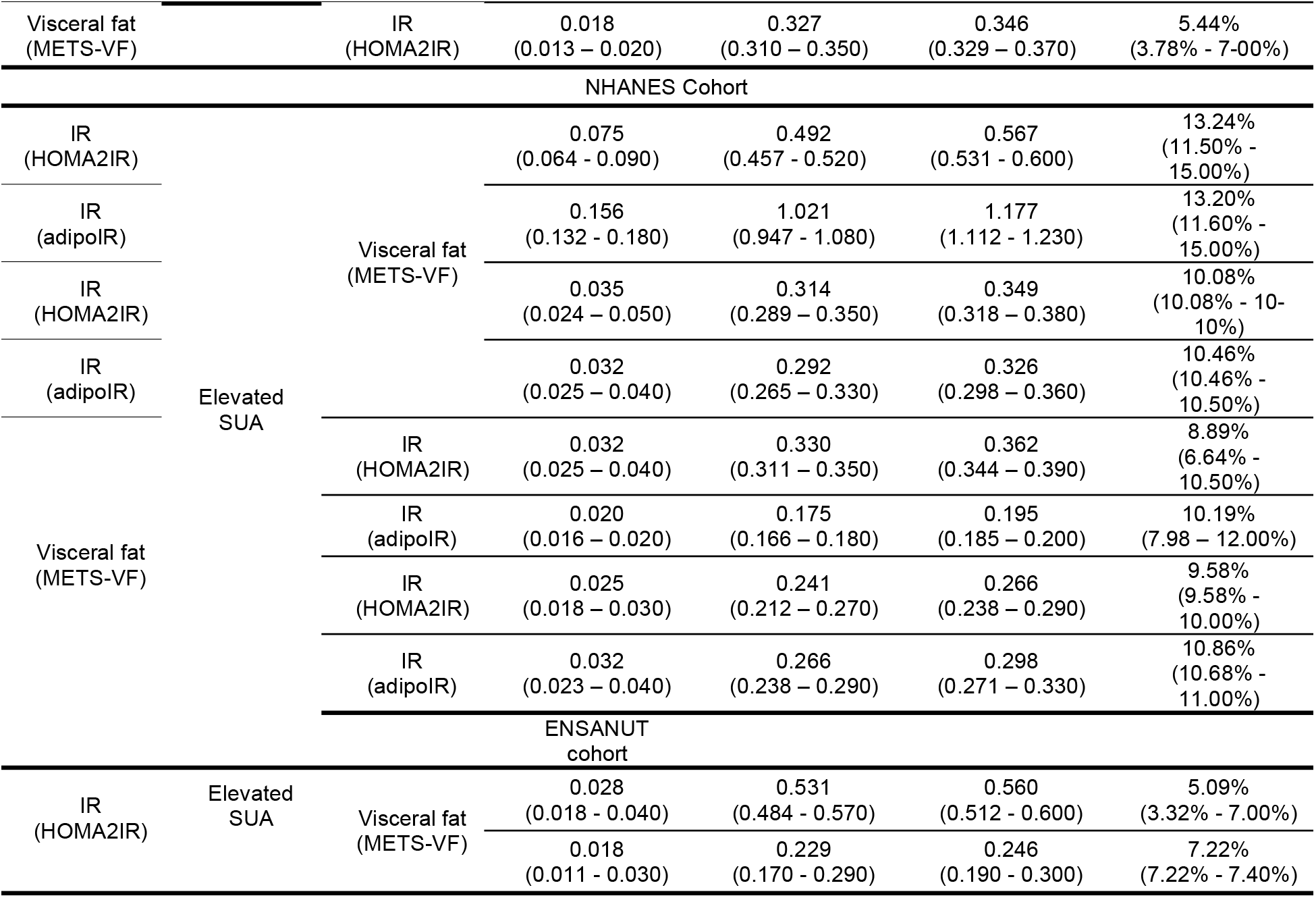
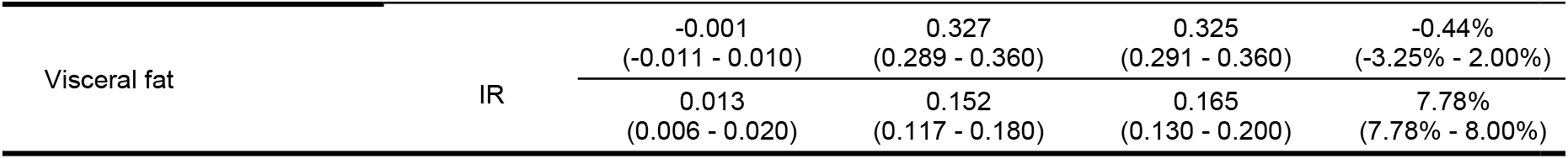
Mediation analyses for all evaluated mechanisms in the UIEM, NHANES, ENSANUT and joint NHANES-ENSANUT cohorts. *Abbreviations*: ACME: Average Causal Mediation Effect. ADE: Average Direct Effect. H2-IR: Homeostatic Model for Insulin Resistance. METS-VF: Metabolic Score for Visceral Fat.

### Adiponectin has a role in the relationship linking elevated SUA, IR, and visceral obesity

Given previous observations, we hypothesized that the impact of elevated SUA on adiponectin levels is likely involved in the identified mediation mechanism^7,20^.Therefore, we investigated the contribution of adiponectin within this mediation framework in the UIEM cohort (**Figure 2B**). A set of 10 mediation models were developed, including two with a joint mediator which accounts for SUA units per unit of serum adiponectin (**Table 2**). When assessing this joint mediator, we observed that the SUA/adiponectin ratio mediated 17.09% (95%CI 8.72-27.00%) of the effect of IR-promoted VAT accumulation and 15.50% (95%CI 5.40-27.00%) of the effect of VAT-promoted peripheral IR (**Supplementary Material**). Notably, both of these proportions were higher than for SUA alone in any direction, indicating that the effect of SUA was likely partly mediated by its effect on adiponectin levels.

### SUA as a clinical marker of IR and VAT accumulation

After confirming the potential mediating role of SUA on peripheral/adipose tissue IR-promoted VAT accumulation, we sought to investigate whether SUA could be used to identify the phenotypes of peripheral or adipose tissue IR or visceral obesity. For our joint cohort, the cutoff value for both phenotypes varied for male (362.83 umol/L for peripheral IR and 321.19 umol/L for visceral obesity), and female sex (321.19 umol/L for peripheral IR and 285.5 umol/L for visceral obesity). None of these models showed SUA to be a strong predictor for its corresponding outcome, as demonstrated by their respective AUROCs, but all displayed high negative predictive values (85.9%, 64.7%, 85.6%, 84.3%, respectively) indicating SUA levels below these cut-off values could be used to rule out peripheral IR and/or visceral obesity (**Supplementary Material**). Therefore, we propose that SUA could be used to potentially rule-out underlying cardiometabolic conditions like IR and visceral obesity in both multiethnic populations.

## DISCUSSION

Here, we proposed a role for elevated SUA levels on the bidirectional relationship between peripheral/adipose tissue IR and VAT accumulation. We built these models under a strong assumption on causality with the aim of assessing the role SUA in cardio-metabolic health. Our research aimed to integrate three known mechanisms: 1) The relationship between peripheral IR and SUA, 2) the role of SUA on VAT accumulation, and 3) the connection between IR and visceral adiposity^3,18,21,22^. Using these relationships, we developed a framework which proposes how elevated SUA links together peripheral and adipose tissue IR with VAT accumulation through a mediating mechanism. Our analyses were first explored using gold standard and anthropometric measurements for peripheral IR and VAT accumulation, while accounting for the involvement of adiponectin to account for adipose tissue dysfunction within the mechanism. Subsequently, we translated these findings to two nationally representative and diverse cohorts to confirm these mechanisms at a population level. Our approach allowed us to develop a more robust and mechanistic characterization of the metabolic pathways underlying the complex interaction of insulin sensitivity with adipose tissue function and distribution and the potential role of elevated SUA in the mechanism. Evidence from previous studies supports the existence of a bidirectional relationship between IR and VAT accumulation, which was confirmed on our model-based causal mediation analyses^23^. We further identified that elevated SUA levels have a larger mediating effect in peripheral/adipose tissue IR-mediated leads to VAT accumulation. Even though the link between SUA levels and peripheral IR in this direction has been the least studied, previous studies have shown that IR may lead to hyperuricemia through an increase in renal resorption of Na^+^ in the proximal tubule and a shift in renal electrolytic balance through the URAT1 transporter^24^. Elevated SUA could lead to VAT accumulation through three known mechanisms: 1) through the increase of uric acid-dependent intracellular and mitochondrial oxidative stress via NADPH oxidase activation, 2) through the inhibition of AMP-activated protein kinase by low intracellular phosphate levels, which decreases transformation rate of AMP to IMP, to inosine and uric acid, and 3) through the activation of the nuclear transcription factor, carbohydrate responsive element-binding protein and increased ketohexokinase expression, responding to fructose abundance and avoiding phosphate depletion^3,25,26^.

A larger body of evidence supports the pathway which indicates that VAT accumulation leads to IR mediated by elevated SUA. Elevated SUA is known to precede IR through different mechanisms, both metabolic and immunological, and VAT accumulation can cause elevated SUA through an increased expression of xanthine oxidoreductase of adipose tissue, commonly found in obesity upstream from PPAR-γ, which is a regulator of adipogenesis^9,27,28^. This could explain our observation of a more important role for elevated SUA as a mediator of VAT accumulation via adipose tissue IR instead of peripheral IR; however, this should be explored in future studies. A hypothesis stating that the temporal causality of this mechanism depends more on previous metabolic states could indicate that the reciprocal effect of IR on VAT accumulation can occur at the same time but not with the same intensity. VAT accumulation via elevated SUA could lead to IR when the subject does not yet have IR, but IR could lead to further VAT accumulation via elevated SUA once the threshold for IR has been met when the process, with possible involvement of additional immuno-metabolic players (**Figure 3**). We considered an non-specific marker for inflammation, C-Reactive Protein, to account for inflammation in our mediation analyses. Nevertheless, neither the association nor the mediations were significant, which was expected given the ambiguity of this biomarker in a specific inflammation context. Further studies are required to evaluate the role of inflammation in modifying the influence of visceral adiposity and insulin resistance on circulating uric acid levels.

**Figure 3.**
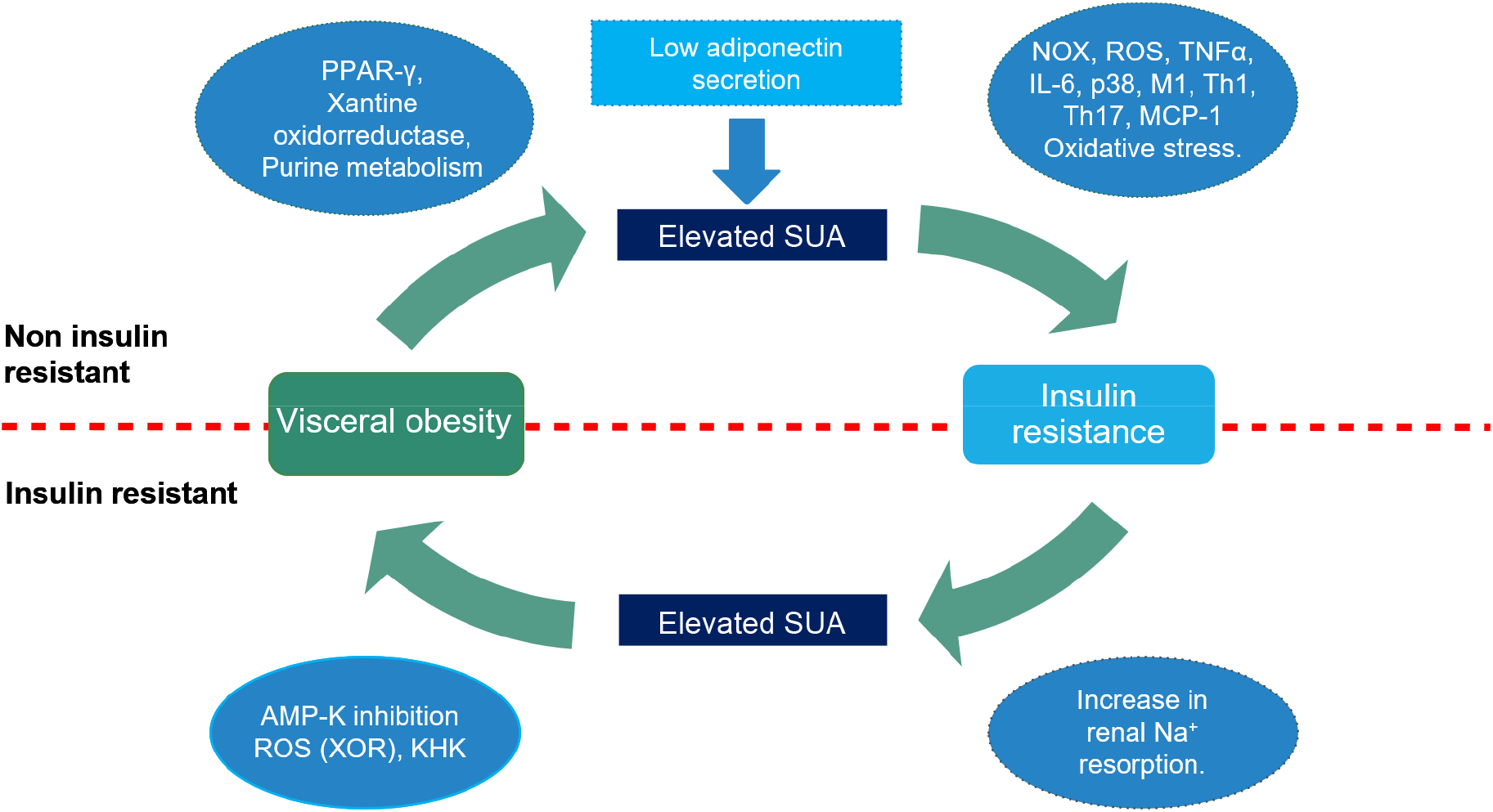
Summary of possible biochemical and immunological causal players which may regulate the influence of serum uric acid, insulin resistance, and visceral adipose tissue accumulation.

As our primary analysis in the UIEM cohort suggests, adiponectin interacts with SUA to modify the bidirectional relationship between VAT and IR. The role of adiponectin in adipose tissue insulin sensitivity and uric acid secretion has been previously described. We identified that all regressions that were adjusted by either eGFR or creatinine improved their goodness-of-fit against those without adjustment. In the context of elevated SUA levels, abundance of adiponectin receptors is increased through an Adipo-R1 pathway, leading to reduced circulating levels. This, together with reduced adiponectin secretion because of visceral fat accumulation, leads to a reduction in the anti-inflammatory of adiponectin, which makes it unable to completely counter the oxidative stress, inflammation, and the change in tissue-specific macrophage profiles caused by elevated SUA levels, leading towards insulin resistance. ^18,29–31^ Our results then suggest that the mediating effect of SUA may be partly mediated by its impact on circulating adiponectin levels.

Elevated SUA has been previously related with cardiometabolic conditions as a biomarker of cardiometabolic health^3,9,22,23^. Despite the relevance of SUA in this setting, a cut-off to identify IR or visceral obesity had not previously been estimated; the observed specificities and sensitivities estimated for SUA in our study cannot support its routine use as a reliable diagnostic tool for either condition, but normal SUA levels can be used to rule out underlying cardio-metabolic phenomena. Although we observed a better performance of SUA in females, further evaluations of underlying hormonal and physiological assessments should be performed to evaluate a potential sexual dimorphism on the impact of SUA on whole-body metabolism. Our study had some strengths and limitations. Strengths of our study include the evaluation of these mechanisms using gold-standard measurements for IR and VAT which also allowed for a characterization of the role of adiponectin within our causality framework. Furthermore, the use of ethnically diverse, representative population-based data, allowed for precise estimates of the relationship between these phenomena at the population level and the investigation of the role of SUA as a biomarker of cardio-metabolic abnormalities.Amongst the limitations to be acknowledged include its cross-sectional nature; although hypothesized, potential causality based on biological plausibility and previous findings but not on temporality of the mechanisms could be established. Furthermore, given the different sources for the data, not all measurements were done the same way accounting for some uncertainty; however, this was addressed considering this variability within a mixed effects framework and the feasibility of combining the cohorts was tested by using PCA to evaluate the impact of different covariance matrices. Lastly, the UIEM cohort was composed of highly selected patients, which limits its external validity, therefore, findings relating adiponectin should be further replicated in a population-based cohort. Overall, our approach allowed the elucidation of a pathophysiological relationship which had not yet been entirely established and which may clarify the role of SUA in cardio-metabolic health.

In conclusion, elevated SUA acts as a mediator inside a bidirectional relationship between IR and VAT accumulation. The role of elevated SUA appears to be larger when considering adipose tissue IR as a promoter of VAT accumulation. Adiponectin appears to be involved as a modifier of the role of elevated SUA within these mechanisms. SUA could be a potential marker to evaluate metabolic health from a pathophysiological and mechanistic perspective. Further longitudinal population-based studies should be performed to clarify the hypothesized temporality and the possible immunometabolic pathways underlying this relationship.

## Data Availability

All data sources and R code are available for reproducibility of results at https://github.com/oyaxbell/hyperuricemia_ir_vat

## ACKNOWLEDGMENTS

AMS, AVV, CAFM, ECG, NEAV are enrolled at the PECEM program of the Faculty of Medicine at UNAM. NEAV, AVV and TLVR and are supported by CONACyT.

## DATA AVAILABILITY

All data sources and R code are available for reproducibility of results at https://github.com/oyaxbell/hyperuricemia_ir_vat, as well as Supplementary Material.

## AUTHOR CONTRIBUTIONS

Research idea and study design LFC, CAAS, OYBC; data acquisition: OYBC, PAV, DGV, TLVR, RR, CAAS; data analysis/interpretation: LFC, OYBC, NEAV, AMS, CAFM, ECG, AVV, PAV, CAAS; statistical analysis: LFC, OYBC; manuscript drafting: LFC, OYBC, CAFM, AMS, ECG, NEAV, AVV, PAV, RR, TLVR, CAAS; supervision or mentorship: OYBC, CAAS. Each author contributed important intellectual content during manuscript drafting or revision and accepts accountability for the overall work by ensuring that questions pertaining to the accuracy or integrity of any portion of the work are appropriately investigated and resolved.

