## Supplementary material for "Elevated serum uric acid is a facilitating mechanism for insulin resistance mediated accumulation of visceral adipose tissue"

**Supplemental Material**

**Supplementary Methods**

a. **UIEM cohort inclusion criteria**

Inclusion criteria for this cohort were as follows:

- SIGMA Cohort study: We recruited healthy subjects without underlying comorbidities and subjects with T2D and glycated hemoglobin (A1c) concentration <8%, who were not receiving insulin and were treated only with metformin. We excluded subjects with active smoking, T2D complications (nephropathy, neuropathy, and retinopathy), cardiovascular diseases, chronic kidney disease, or with an acute infection from all analyses.
- Cohort of non-diabetic patients: Subjects with either obesity (BMI≥ 40kg/m2) or normal weight (BMI<25kg/m2) were included. Those with ≤ 1 NCEP ATPIII criteria for metabolic syndrome (except for waist circumference) were considered as metabolically-healthy and those with ≥2 as metabolically-unhealthy patients. We excluded subjects with acute infection, chronic hepatic disease, autoimmune diseases, alcohol consumption >10g/day, active smoking, fasting triglycerides >500mg/dL and height >150cm.

b. **Logistic models and elevated SUA cut-offs**

There is currently no consensus for established cut-offs to determine elevated SUA levels in relation to cardio metabolic disorders, and population-based studies have identified different cut-off values depending on the underlying phenotype under study (29–31). To overcome this limitation, we established sex-specific cutoffs taking into consideration a midpoint between hypouricemia (32,33). and serum solubility, (3) thus weighing its J-shaped antioxidant behavior. For males the cut-off was established at SUA>6.0 mg/dL (356.88 µmol/L), and for females at SUA>4.8 mg/dL (285.5 µmol/L). These cutoffs were used to carry out all phenotype-related models.

To evaluate the phenotypes of IR, visceral obesity and elevated SUA using dichotomized predictors, we fitted multiple logistic regression models independently for ENSANUT and NHANES and for the joint cohort, where models were specified with cohort of origin as a random intercept to account for clustering. These models were also fitted by age, sex, ethnicity (only for NHANES and joint cohorts), and serum creatinine or eGFR wherever appropriate. Goodness of fit was assessed in logistic regression models using the Hosmer-Lemeshow test and models were selected using BIC minimization.

**Supplementary Tables**

|  | General population | Normal SUA | Elevated SUA | *p*-values |
| --- | --- | --- | --- | --- |
| n | 4402 | 2304 | 2098 |  |
| Age [years] | 47.00 (33.00) | 43.00 (32.00) | 51.00 (32.00) | <0.001 |
| Female sex [n,%] | 2160 (49.07%) | 1061 (46.05%) | 1099 (52.38%) | **0.414** |
| Glucose [mmol/L] | 5.43 (0.90) | 5.33 (0.83) | 5.55 (1.00) | <0.001 |
| Insulin [pmol/L] | 56.72 (57.66) | 46.20 (45.42) | 67.08 (66.99) | <0.001 |
| Uric acid [µmol/L] | 315.24 (113.01) | 267.66 (77.32) | 380.67 (83.27) | <0.001 |
| Antihiperuricemics [n,%] | 11 (0.25%) | 5 (0.22%) | 6 (0.29%) | **0.763** |
| TGL [mmol/L] | 1.22 (0.94) | 1.08 (0.81) | 1.39 (1.01) | <0.001 |
| cHDL [mmol/L] | 1.32 (0.49) | 1.37 (0.52) | 1.27 (0.47) | <0.001 |
| Total cholesterol [mmol/L] | 4.95 (1.42) | 4.89 (1.37) | 5.00 (1.42) | <0.001 |
| BMI [kg/m^2^] | 27.44 (7.91) | 25.87 (7.02) | 29.17 (8.01) | <0.001 |
| WHtR | 0.58 (0.13) | 0.55 (0.12) | 0.60 (0.13) | <0.001 |
| Palmitate [µmol/L] | 2690.00  (1140.00) | 2550.00 (1032.50) | 2850.00 (1230.00) | <0.001 |
| Creatinine [mmol/L] | 74.72 (26.52) | 70.72 (22.10) | 74.26 (24.75) | <0.001 |
| eGFR[mL/min/1.73m^2^] | 97.67 (33.39) | 103.09 (30.46) | 91.60 (35.63) | <0.001 |
| HOMA2%B | 79.20 (52.00) | 74.00 (46.20) | 86.55 (55.18) | <0.001 |
| HOMA2%S | 94.90 (95.00) | 112.60 (108.90) | 77.05 (77.45) | <0.001 |
| HOMA2IR | 1.05 (1.12) | 0.89 (0.90) | 1.30 (1.30) | <0.001 |
| Insulin resistant [HOMA2IR] | 879 (19.97%) | 305 (13.24%) | 574 (27.36%) | <0.001 |
| METS-VF | 6.92 (1.01) | 6.68 (1.14) | 7.12 (0.75) | <0.001 |
| People with visceral obesity [METS-VF] | 1414 (32.12%) | 505 (21.92%) | 909 (43.33%) | *<*0.001 |
| Subjects with diabetes [n,%] | 241 (5.47%) | 83 (3.60%) | 158 (7.53%) | *<*0.001 |
| Hypertensives [n,%] | 745 (16.92%) | 327 (14.19%) | 418 (19.92%) | 0.001 |
| Antihypertensives [n,%] | 1495 (33.96%) | 579 (25.13%) | 916 (43.66%) | *<*0.001 |
| Aspirin [n,%] | 14 (0.32%) | 4 (0.17%) | 10 (0.48%) | **0.109** |
| Hypercholesterolemia [n,%] | 1371 (31.14%) | 628 (27.26%) | 743 (35.41%) | 0.002 |
| Antihyperlipidemics [n,%] | 388 (8.81%) | 200 (8.68%) | 188 (8.96%) | **0.542** |
| Mexican-Americans | 657 (14.93%) | 385 (16.71%) | 272 (12.96%) | *<*0.001 |
| Non-hispanic White [n,%] | 1 968 (44.71%) | 979 (42.49%) | 989 (47.14%) | **0.822** |
| Non-hispanic Black [n,%] | 928 (21.08%) | 467 (20.27%) | 461 (21.97%) | **0.844** |
| Other ethnicity [n,%] | 849 (19.29%) | 473 (20.53%) | 376 (17.92%) | 0.001 |

**Supplementary Table 1.** General characteristics of studied cohort (NHANES). Patients with elevated SUA are those with a serum concentration greater than 288.48 umol/L for females and 356.88 umol/L for males. *Abbreviations*: BMI: Body Mass Index. cHDL: High Density Lipoprotein cholesterol. HOMA2IR: Homeostatic Model for Insulin Resistance. HOMA2%S: Homeostatic Model for Insulin Resistance for pancreatic β cell sensitivity. HOMA2%B: Homeostatic Model for Insulin Resistance for functionality of pancreatic β cells. METS-VF: Metabolic Score for Visceral Fat. WHtR: Waist-height ratio.

|  | General population | Normal SUA | Elevated SUA | *p-*values |
| --- | --- | --- | --- | --- |
| n | 1914 | 906 | 1008 |  |
| Age [years] | 45.00 (24.00) | 44.00 (24.00) | 46.00 (25.00) | 0.081 |
| Female sex [n,%] | 728 (38.04%) | 327 (36.09%) | 401 (39.78%) | 0.006 |
| Glucose [mmol/L] | 5.32 (0.97) | 5.22 (0.94) | 5.38 (0.94) | 0.001 |
| Insulin [pmol/L] | 60.42 (58.34) | 50.00 (49.13) | 72.23 (67.71) | <0.001 |
| Uric acid [µmol/L] | 315.24 (113.01) | 261.71 (49.13) | 368.78 (67.71) | <0.001 |
| TGL [mmol/L] | 1.90 (1.45) | 1.69 (1.46) | 2.06 (1.41) | <0.001 |
| cHDL [mmol/L] | 0.96 (0.31) | 0.98 (0.31) | 0.93 (0.28) | <0.001 |
| Total cholesterol [mmol/L] | 4.79 (1.29) | 4.71 (1.24) | 4.87 (1.34) | <0.001 |
| BMI [kg/m^2^] | 27.99 (6.71) | 26.58 (6.37) | 29.24 (6.49) | <0.001 |
| WHtR | 0.60 (0.11) | 0.59 (0.11) | 0.62 (0.11) | <0.001 |
| Creatinine [mmol/L] | 61.00 (21.22) | 58.34 (16.80) | 64.53 (22.98) | <0.001 |
| eGFR [mL/min/1.73m^2^] | 107.01 (23.11) | 109.49 (18.99) | 103.63 (27.26) | <0.001 |
| HOMA2%B | 86.05 (61.58) | 76.20 (56.70) | 95.20 (59.50) | <0.001 |
| HOMA2%S | 83.20 (86.20) | 102.50 (100.70) | 72.40 (71.90) | <0.001 |
| HOMA2IR | 1.20 (1.18) | 0.98 (1.03) | 1.38 (1.31) | <0.001 |
| Insulin resistant  [HOMA2IR] | 382 (19.96%) | 131 (14.46%) | 251 (24.90%) | *<*0.001 |
| Subjects with  diabetes [n,%] | 101 (5.28%) | 41 (4.53%) | 60 (5.95%) | 0.059 |
| METS-VF | 7.07 (0.67) | 6.93 (0.82) | 7.17 (0.59) | *<*0.001 |
| People with visceral obesity [METS-VF] | 759 (39.66%) | 292 (32.23%) | 467 (46.33%) | *<*0.001 |

**Supplementary table 2.** General characteristics of ENSANUT cohort. Patients with elevated SUA are those with a serum concentration greater than 288.48 umol/L for females and 356.88 umol/L for males. *Abbreviations:* BMI: Body Mass Index. cHDL: High Density Lipoprotein cholesterol. HOMA2IR: Homeostatic Model for Insulin Resistance. HOMA2%S: Homeostatic Model for Insulin Resistance for pancreatic β cell sensitivity. HOMA2%B: Homeostatic Model for Insulin Resistance for functionality of pancreatic β cells. Insulin resistance. METS-VF: Metabolic Score for Visceral Fat. WHtR: Waist-height ratio.

| Model | Parameter | Beta | SE | p-value | 95% CI |
| --- | --- | --- | --- | --- | --- |
| HOMA2IR ~ METS-VF | METS-VF | 28.865 | 0.761 | <0.001 | 27.372 - 30.359 |
|  | METS-VF^2^ | 1.979 | 0.666 | 0.003 | 0.674 - 3.285 |
|  | METS-VF^3^ | -4.415 | 0.662 | <0.001 | -5.712 - -3.118 |
| SUA ~ METS-VF | METS-VF | 0.101 | 0.004 | <0.001 | 0.093 - 0.109 |
| HOMA2IR ~ SUA | SUA | 0.827 | 0.043 | <0.001 | 0.743 - 0.911 |
| METSVF ~ HOMA2IR | HOMA2IR | 28.788 | 0.761 | <0.001 | 27.296 - 30.28 |
|  | HOMA2IR^2^ | 3.004 | 0.801 | <0.001 | 1.426 - 4.583 |
|  | HOMA2IR^3^ | -5.621 | 0.816 | <0.001 | -7.234 - -4.007 |
| SUA ~ HOMA2IR | HOMA2IR | 0.094 | 0.005 | <0.001 | 0.085 - 0.104 |
| METS-VF ~ SUA | SUA | 16.333 | 0.880 | <0.001 | 14.608 - 18.058 |
|  | SUA^2^ | 1.351 | 0.837 | 0.11 | -0.289 - 2.991 |
|  | SUA^3^ | -3.327 | 0.833 | <0.001 | -4.961 - -1.693 |
| HOMA2IR ~ METS-VF + SUA | METS-VF | 26.951 | 0.784 | <0.001 | 25.414 - 28.489 |
|  | METS-VF^2^ | 1.829 | 0.660 | 0.0056 | 0.535 - 3.123 |
|  | METS-VF^3^ | -4.160 | 0.656 | <0.001 | -5.446 - -2.874 |
|  | SUA | 0.323 | 0.035 | <0.001 | 0.254 - 0.392 |
| SUA ~ HOMA2IR + METS-VF | METS-VF | 0.060 | 0.006 | <0.001 | 0.047 - 0.072 |
|  | HOMA2IR | 0.064 | 0.006 | <0.001 | 0.053 - 0.075 |
| METS-VF ~ SUA + HOMA2IR | SUA | 8.980 | 0.802 | <0.001 | 7.407 - 10.553 |
|  | SUA^2^ | 0.068 | 0.739 | 0.93 | -1.381 - 1.518 |
|  | SUA^3^ | -3.137 | 0.737 | <0.001 | -4.582 - -1.692 |
|  | HOMA2IR | 26.969 | 0.771 | <0.001 | 25.457 - 28.48 |
|  | HOMA2IR^2^ | 2.763 | 0.792 | <0.001 | 1.201 - 4.325 |
|  | HOMA2IR^3^ | -5.336 | 0.807 | <0.001 | -6.931 - -3.74 |
| adipoIR ~ METS-VF | METS-VF | 15.196 | 0.386 | <0.001 | 14.439 - 15.952 |
|  | METS-VF^2^ | 0.012 | 0.339 | 0.97 | -0.652 - 0.675 |
|  | METS-VF^3^ | -1.997 | 0.338 | <0.001 | -2.659 - -1.334 |
| adipoIR ~ SUA | SUA | 9.330 | 0.426 | <0.001 | 8.495 - 10.164 |
|  | SUA^2^ | 1.311 | 0.371 | <0.001 | 0.582 - 2.039 |
| METSVF ~ adipoIR | adipoIR | 29.406 | 0.751 | <0.001 | 27.933 - 30.879 |
|  | adipoIR^2^ | 0.543 | 0.774 | 0.48 | -0.977 - 2.064 |
|  | adipoIR^3^ | -4.926 | 0.791 | <0.001 | -6.484 - -3.368 |
| SUA ~ adipoIR | adipoIR | 0.214 | 0.010 | <0.001 | 0.195 - 0.233 |
| METS-VF ~ SUA + adipoIR | SUA | 8.231 | 0.805 | <0.001 | 6.653 - 9.81 |
|  | SUA^2^ | -0.366 | 0.740 | 0.62 | -1.816 - 1.085 |
|  | SUA^3^ | -3.191 | 0.732 | <0.001 | -4.627 - -1.756 |
|  | adipoIR | 27.261 | 0.773 | <0.001 | 25.746 - 28.777 |
|  | adipoIR^2^ | 0.410 | 0.766 | 0.59 | -1.095 - 1.916 |
|  | adipoIR^3^ | -4.617 | 0.783 | <0.001 | -6.159 - -3.075 |
| SUA ~ METS-VF + adipoIR | SUA | 0.059 | 0.006 | <0.001 | 0.048 - 0.07 |
|  | SUA^2^ | 0.136 | 0.012 | <0.001 | 0.112 - 0.161 |
| adipoIR ~ METS-VF + SUA | METS-VF | 14.015 | 0.396 | <0.001 | 13.24 - 14.791 |
|  | METS-VF^2^ | -0.139 | 0.334 | 0.68 | -0.795 - 0.517 |
|  | METS-VF^3^ | -1.861 | 0.333 | <0.001 | -2.514 - -1.208 |
|  | SUA | 3.954 | 0.362 | <0.001 | 3.243 - 4.664 |
|  | SUA^2^ | 1.191 | 0.331 | <0.001 | 0.543 - 1.84 |

**Supplementary table 3.** Regression analysis coefficients for NHANES cohort. All models were adjusted by sex, age, ethnicity, and either serum creatinine or eGFR where appropriate. *Abbreviations:* adipoIR: Adipose Tissue Insulin Resistance. HOMA2IR: Homeostatic Model for Insulin Resistance. METS-VF: Metabolic Score for Visceral Fat. SUA: serum uric acid.

| Outcome | Predictor | OR | SE | p-value | 95% CI |
| --- | --- | --- | --- | --- | --- |
| Elevated METS-VF | Elevated HOMAIR | 5.567 | 0.088 | <0.001 | 4.681 – 6.619 |
| Elevated HOMA2IR | Elevated METS-VF | 5.662 | 0.090 | <0.001 | 4.748 - 6.75 |
| Elevated METS-VF | Elevated SUA | 2.132 | 0.072 | <0.001 | 1.852 - 2.454 |
| Elevated SUA | Elevated METS-VF | 2.116 | 0.072 | <0.001 | 1.838 - 2.436 |
| Elevated HOMA2IR | Elevated SUA | 2.588 | 0.081 | <0.001 | 2.208 - 3.033 |
| Elevated SUA | Elevated HOMAIR | 2.587 | 0.082 | <0.001 | 2.203 - 3.039 |
| Elevated SUA | Joint variable | 1.468 | 0.030 | <0.001 | 1.385 - 1.557 |
| Elevated METS-VF | Elevated adipoIR | 5.022 | 0.081 | <0.001 | 4.282 - 5.889 |
| Elevated adipoIR | Elevated METS-VF | 5.095 | 0.082 | <0.001 | 4.335 - 5.989 |
| Elevated adipoIR | Elevated SUA | 2.689 | 0.075 | <0.001 | 2.323 - 3.112 |
| Elevated SUA | Elevated adipoIR | 2.669 | 0.075 | <0.001 | 2.303 - 3.094 |

**Supplementary table 4.** Logistic regression analysis coefficients for NHANES cohort. All models were adjusted by sex, age, ethnicity, and either serum creatinine or eGFR where appropriate. Elevated cutoffs were established as described in methods section. Joint variable refers to IR and METS-VF joint categorization. *Abbreviations:* adipoIR: Adipose Tissue Insulin Resistance. HOMA2IR: Homeostatic Model for Insulin Resistance. METS- VF: Metabolic Score for Visceral Fat. SUA: serum uric acid.

| Model | Parameter | Beta | SE | p-value | 95% CI |
| --- | --- | --- | --- | --- | --- |
| HOMA2IR ~ METS-VF | METS-VF | 0.380 | 0.017 | <0.001 | 0.346 - 0.413 |
| SUA ~ METS-VF | METS-VF | 0.070 | 0.006 | <0.001 | 0.058 - 0.082 |
| HOMA2IR ~ SUA | SUA | 0.452 | 0.065 | <0.001 | 0.324 - 0.58 |
| METSVF ~ HOMA2IR | HOMA2IR | 0.567 | 0.015 | <0.001 | 0.537 - 0.597 |
| SUA ~ HOMA2IR | HOMA2IR | 0.094 | 0.005 | <0.001 | 0.085 - 0.104 |
| METS-VF ~ SUA | SUA | 0.807 | 0.044 | <0.001 | 0.721 - 0.892 |
| HOMA2IR ~ METS-VF + SUA | METS-VF | 0.408 | 0.012 | <0.001 | 0.384 - 0.431 |
|  | SUA | 0.335 | 0.035 | <0.001 | 0.266 - 0.405 |
| SUA ~ HOMA2IR + METS-VF | METS-VF | 0.060 | 0.006 | <0.001 | 0.047 - 0.072 |
|  | HOMA2IR | 0.064 | 0.006 | <0.001 | 0.053 - 0.075 |
| METSVF ~ SUA + HOMA2IR | SUA | 0.460 | 0.040 | <0.001 | 0.382 - 0.539 |
|  | HOMA2IR | 0.530 | 0.015 | <0.001 | 0.5 - 0.56 |

**Supplementary table 5.** Regression analysis coefficients for ENSANUT cohort. All models were adjusted by sex, age, and either serum creatinine or eGFR where appropriate. *Abbreviations:* HOMA2IR: Homeostatic Model for Insulin Resistance. METS-VF: Metabolic Score for Visceral Fat. SUA: serum uric acid.

| Outcome | Predictor | OR | SE | p-value | 95% CI |
| --- | --- | --- | --- | --- | --- |
| Elevated METS-VF | Elevated HOMA2IR | 3.269 | 0.125 | <0.001 | 2.559 - 4.175 |
| Elevated HOMA2IR | Elevated METS-VF | 3.302 | 0.126 | <0.001 | 2.58 - 4.226 |
| Elevated METS-VF | Elevated SUA | 1.440 | 0.100 | <0.001 | 1.182 - 1.753 |
| Elevated SUA | Elevated METS-VF | 1.642 | 0.098 | <0.001 | 1.354 - 1.99 |
| Elevated HOMA2IR | Elevated SUA | 2.082 | 0.123 | <0.001 | 1.637 - 2.648 |
| Elevated SUA | Elevated HOMA2IR | 2.063 | 0.124 | <0.001 | 1.618 - 2.63 |
| Elevated SUA | Joint variable | 1.322 | 0.043 | <0.001 | 1.216 - 1.438 |

**Supplementary table 6**. Logistic regression analysis coefficients for ENSANUT cohort. All models were adjusted by sex, age, and either serum creatinine or eGFR where appropriate. Elevated cutoffs were established as described in Methods section. Joint variable refers to IR and METS-VF joint categorization. *Abbreviations:* HOMA2IR: Homeostatic Model for Insulin Resistance. METS-VF: Metabolic Score for Visceral Fat. SUA: serum uric acid.

| Model | Parameter | Beta | SE | p-value | 95% CI |
| --- | --- | --- | --- | --- | --- |
| HOMA2IR ~ METS-VF | METS-VF | 32.915 | 0.750 | <0.001 | 31.445 - 34.385 |
|  | METS-VF^2^ | 0.853 | 0.669 | 0.2 | -0.46 - 2.165 |
|  | METS-VF^3^ | -5.013 | 0.665 | <0.001 | -6.316 - -3.709 |
| SUA ~ METS-VF | METS-VF | 0.092 | 0.004 | <0.001 | 0.085 - 0.099 |
| HOMA2IR ~ SUA | SUA | 0.511 | 0.032 | <0.001 | 0.447 - 0.574 |
| METSVF ~ HOMA2IR | HOMA2IR | 35.108 | 0.873 | <0.001 | 33.396 - 36.82 |
|  | HOMA2IR^2^ | 1.843 | 0.873 | 0.035 | 0.131 - 3.556 |
|  | HOMA2IR^3^ | -6.636 | 0.886 | <0.001 | -8.375 - -4.897 |
| SUA ~ HOMA2IR | HOMA2IR | 4.499 | 0.285 | <0.001 | 3.941 - 5.058 |
|  | HOMA2IR^2^ | 0.287 | 0.283 | 0.31 | -0.268 - 0.842 |
|  | HOMA2IR^3^ | -1.334 | 0.283 | <0.001 | -1.889 - -0.779 |
| METS-VF ~ SUA | SUA | 18.463 | 0.926 | <0.001 | 16.646 - 20.28 |
|  | SUA^2^ | 1.327 | 0.863 | 0.12 | -0.364 - 3.018 |
|  | SUA^3^ | -4.039 | 0.870 | <0.001 | -5.746 - -2.332 |
| HOMA2IR ~ METS-VF + SUA | METS-VF | 31.894 | 0.778 | <0.001 | 30.369 - 33.419 |
|  | METS-VF^2^ | 0.591 | 0.664 | 0.37 | -0.712 - 1.893 |
|  | METS-VF^3^ | -4.762 | 0.661 | <0.001 | -6.058 - -3.467 |
|  | SUA | 0.200 | 0.030 | <0.001 | 0.141 - 0.258 |
| SUA ~ HOMA2IR + METS-VF | METS-VF | 0.032 | 0.005 | <0.001 | 0.022 - 0.042 |
|  | HOMA2IR | 0.081 | 0.004 | <0.001 | 0.073 - 0.089 |
| METS-VF ~ SUA + HOMA2IR | SUA | 11.538 | 0.844 | <0.001 | 9.879 - 13.196 |
|  | SUA^2^ | 0.708 | 0.766 | 0.355 | -0.793 - 2.209 |
|  | SUA^3^ | -3.016 | 0.774 | <0.001 | -4.534 - -1.498 |
|  | HOMA2IR | 32.018 | 0.789 | <0.001 | 30.47 - 33.566 |
|  | HOMA2IR^2^ | 1.809 | 0.772 | 0.019 | 0.295 - 3.322 |
|  | HOMA2IR^3^ | -5.937 | 0.789 | <0.001 | -7.488 - -4.386 |

**Supplementary table 7**. Regression analysis coefficients for joint cohort. All models were adjusted by sex, age, ethnicity, and either serum creatinine or eGFR where appropriate. *Abbreviations:* HOMA2IR: Homeostatic Model for Insulin Resistance. METS-VF: Metabolic Score for Visceral Fat. SUA: serum uric acid.

| Outcome | Predictor | OR | SE | p-value | 95% CI |
| --- | --- | --- | --- | --- | --- |
| Elevated METS-VF | Elevated HOMA2IR | 4.682 | 0.072 | <0.001 | 4.064 - 5.393 |
| Elevated HOMA2IR | Elevated METS-VF | 4.742 | 0.073 | <0.001 | 4.109 - 5.473 |
| Elevated METS-VF | Elevated SUA | 1.868 | 0.058 | <0.001 | 1.666 - 2.095 |
| Elevated SUA | Elevated METS-VF | 1.860 | 0.058 | <0.001 | 1.659 - 2.085 |
| Elevated HOMA2IR | Elevated SUA | 2.426 | 0.068 | <0.001 | 2.125 - 2.769 |
| Elevated SUA | Elevated HOMA2IR | 2.415 | 0.068 | <0.001 | 2.112 - 2.761 |
| Elevated SUA | Joint variable | 1.463 | 0.024 | <0.001 | 1.396 - 1.534 |

**Supplementary Table 8.** Logistic regression analysis coefficients for joint cohort. All models were adjusted by sex, age, ethnicity, and either serum creatinine or eGFR where appropriate. Elevated cutoffs were established as described in Methods section. Joint variable refers to IR and METS-VF joint categorization. *Abbreviations:* HOMA2IR: Homeostatic Model for Insulin Resistance. METS-VF: Metabolic Score for Visceral Fat. SUA: serum uric acid.

| **Causality model** | **Efector** | **Mediator** | **Result** | **ACME** | **ADE** | **Total effect** | **% Mediated** |
| --- | --- | --- | --- | --- | --- | --- | --- |
| H2-IR*_log_* | IR  (HOMA2IR) | Elevated SUA | Visceral fat  (METS-VF) | 0.028  (0.022 – 0.040) | 0.289  (0.259 – 0.330) | 0.318  (0.289 – 0.350) | 8.89%  (8.89% - 9.00%) |
| H2-IR’*_log_* | Visceral fat  (METS-VF) |  | IR (HOMA2IR) | 0.028  (0.022– 0.040) | 0.293  (0.263 – 0.330) | 0.321  (0.293 – 0.350) | 8.72%  (8.72-9.00%) |

**Supplementary table 9**. Logistic mediation analyses for general bidirectional mechanisms in joint cohort. *Abbreviations*: ACME: Average Causal Mediation Effect. ADE: Average Direct Effect. AdipoIR: Adipose Insulin Resistance index. H2-IR: Homeostatic Model for Insulin Resistance. METS-VF: Metabolic Score for Visceral Fat.

| **Parameter** | **AUC** | **Cut-off** | **Se** | **Sp** | **PPV** | **NPV** |
| --- | --- | --- | --- | --- | --- | --- |
| IR (Male)  (HOMA2IR) | 0.611  (0.580 - 0.642) | 362.83 umol/L | 61.9%  (57.1% - 66.5%) | 56.3%  (54.0% - 58.7%) | 26.1%  (24.2% - 30.1%) | 85.6%  (83.0% - 86.7%) |
| IR (Female)  (HOMA2IR) | 0.630  (0.609 – 0.651) | 321.19 umol/L | 51.7%  (46.9% - 56.4%) | 76.5%  (74.5% - 78.5%) | 35.5%  (33.0% - 40.0%) | 86.3%  (83.9% - 87.6%) |
| IR (Male)  (adipoIR) | 0.626  (0.599 – 0.654) | 362.83 umol/L | 61.5%  (57.3% – 65.6%) | 57.5%  (55.1% - 60.0%) | 32.9%  (30.7% - 36.9%) | 81.6%  (78.8% - 83.0%) |
| IR (Female)  (adipoIR) | 0.688  (0.662 – 0.714) | 297.4 umol/L | 62.6%  (58.4% - 66.6%) | 65.2%  (62.8% - 67.5%) | 37.2%  (34.9% - 41.4%) | 84.1%  (81.6% - 85.4%) |
| Visceral obesity (Male)  (METS-VF) | 0.598  (0.573 – 0.622) | 321.19 umol/L | 60.1%  (56.7% - 63.4%) | 55.2%  (52.4% - 57.9%) | 46.8%  (44.0% - 50.3%) | 67.9%  (64.8% - 70.2%) |
| Visceral obesity (Female)  (METS-VF) | 0.722  (0.698 – 0.747) | 285.5 umol/L | 74.6%  (70.8% - 78.2%) | 61.1%  (58.7%– 63.4%) | 38.9%  (36.6% - 43.7%) | 87.9%  (85.6% - 88.9%) |

**Supplementary table 10.** ROC curves estimated with categorized variables in NHANES population against SUA as a continuous predictor. *Abbreviations*: AdipoIR: Adipose Insulin Resistance index. AUC: Area under curve. HOMA2IR: Homeostatic Model for Insulin Resistance. IR: Insulin resistance. METS-VF: Metabolic Score for Visceral Fat. NPV: Negative predictive value. PPV: Positive predictive value. Se: Sensibility. Sp: Specificity.

| **Parameter** | **AUC** | **Cut-off** | **Se** | **Sp** | **PPV** | **NPV** |
| --- | --- | --- | --- | --- | --- | --- |
| IR (Male)  (HOMA2IR) | 0.575  (0.520 – 0.631) | 285.5 umol/L | 62.4%  (53.0% - 71.2%) | 52.2%  (48.2% - 56.2%) | 19.8%  (17.4% - 26.9%) | 88.0%  (83.2% - 89.6%) |
| IR (Female)  (HOMA2IR) | 0.608  (0.570 – 0.647) | 303.35 umol/L | 60.0%  (53.8% - 65.9%) | 59.1%  (55.8% - 62.2%) | 29.3%  (26.7% - 34.9%) | 83.9%  (80.2% - 85.2%) |
| Visceral obesity (Male)  (METS-VF) | 0.558  (0.517 – 0.600) | 434.2 umol/L | 27.3%  (22.7% - 32.2%) | 84.0%  (79.9% - 87.6%) | 62.0%  (55.3% - 67.4%) | 54.7%  (48.6% - 61.8%) |
| Visceral obesity (Female)  (METS-VF) | 0.618  (0.584 – 0.652) | 285.5 umol/L | 67.7%  (62.9% - 72.3%) | 52.4%  (48.9% - 55.9%) | 41.6%  (38.2% - 46.9%) | 76.5%  (72.4% - 78.9%) |

**Supplementary table 11.** ROC curves estimated with categorized variables in ENSANUT population against SUA as a continuous predictor. *Abbreviations:* AdipoIR: Adipose Insulin Resistance index. AUC: Area under curve. HOMA2IR: Homeostatic Model for Insulin Resistance. IR: Insulin resistance. METS-VF: Metabolic Score for Visceral Fat. NPV: Negative predictive value. PPV: Positive predictive value. Se: Sensibility. Sp: Specificity.

| **Parameter** | **AUC** | **Cut-off** | **Se** | **Sp** | **PPV** | **NPV** |
| --- | --- | --- | --- | --- | --- | --- |
| IR (Male)  (HOMA2IR) | 0.602  (0.575 - 0.629) | 6.2 | 59.2%  (55.0% - 63.4%) | 57.8%  (55.8% - 59.8%) | 24.6%  (23.2% - 28.1%) | 85.9%  (83.6% - 86.9%) |
| IR (Female)  (HOMA2IR) | 0.657  (0.634 – 0.680) | 5.2 | 56.4%  (52.7% - 60.1%) | 68.0%  (66.3% - 69.8%) | 31.6%  (29.9% - 35.0%) | 85.6%  (83.7% - 86.6%) |
| Visceral obesity (Male)  (METS-VF) | 0.589  (0.568 – 0.610) | 6.0 | 59.5%  (56.7% - 62.3%) | 53.7%  (51.3% - 56.1%) | 48.2%  (45.7% - 51.1%) | 64.7%  (62.1% - 66.9%) |
| Visceral obesity (Female)  (METS-VF) | 0.686  (0.666 – 0.706) | 4.8 | 71.7%  (68.8% - 74.6%) | 58.5%  (56.5% - 60.4%) | 40-0%  (38.1% - 43.5%) | 84.3%  (82.3% - 85.3%) |

**Supplementary table 12.** ROC curves estimated with categorized variables in joint ENSANUT-NHANES population against SUA as a continuous predictor. *Abbreviations:* AdipoIR: Adipose Insulin Resistance index. AUC: Area under curve. HOMA2IR: Homeostatic Model for Insulin Resistance. IR: Insulin resistance. METS-VF: Metabolic Score for Visceral Fat. NPV: Negative predictive value. PPV: Positive predictive value. Se: Sensibility. Sp: Specificity

**Supplementary Figures**

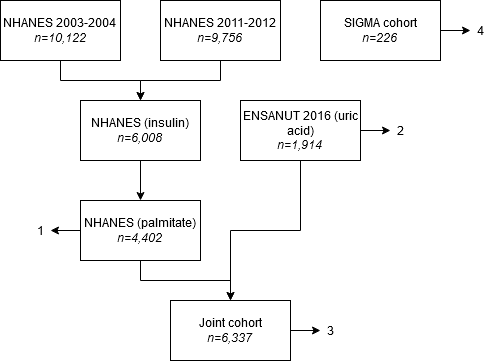

**Supplementary Figure 1:** Flowchart illustrating sample size. For NHANES analysis (1), subjects with insulin and palmitate measurements were selected. For ENSANUT analysis (2), subjects with uric acid measurements were selected. For joint analysis, subjects from 1 and 2 were analyzed together (3). For gold-standard analysis (4), UIEM cohort was used.

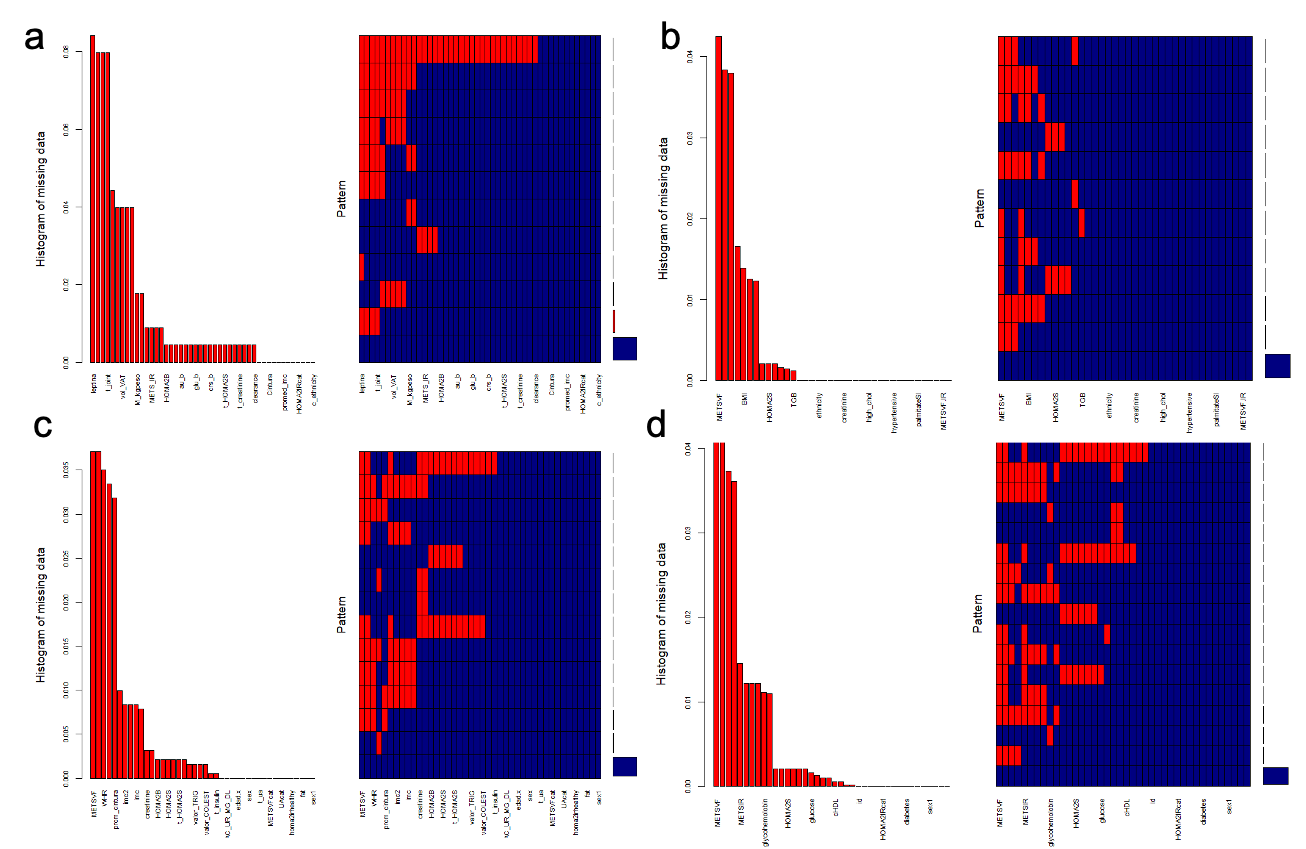

**Supplementary Figure 2:** Missing data histograms for variables in different cohorts. (a) NHANES-ENSANUT joint cohort. (b) NHANES cohort. (c) ENSANUT cohort. (d) Confirmatory cohort. Abbreviations: AdipoIR: Adipose Insulin Resistance index. HOMA2-IR: Homeostatic Model for Insulin Resistance. HOMA2%S: Homeostatic Model for Insulin Resistance for pancreatic β cell sensitivity. HOMA2%B: Homeostatic Modelfor Insulin Resistance for functionality of pancreatic β cells. METS-VF: Metabolic Score for Visceral Fat.

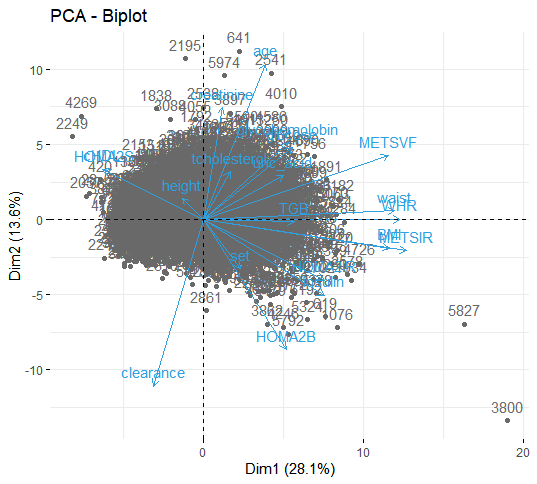

**Supplementary Figure 3:** Biplot illustrating PCA analysis to allow the union of NHANES and ENSANUTcohorts.

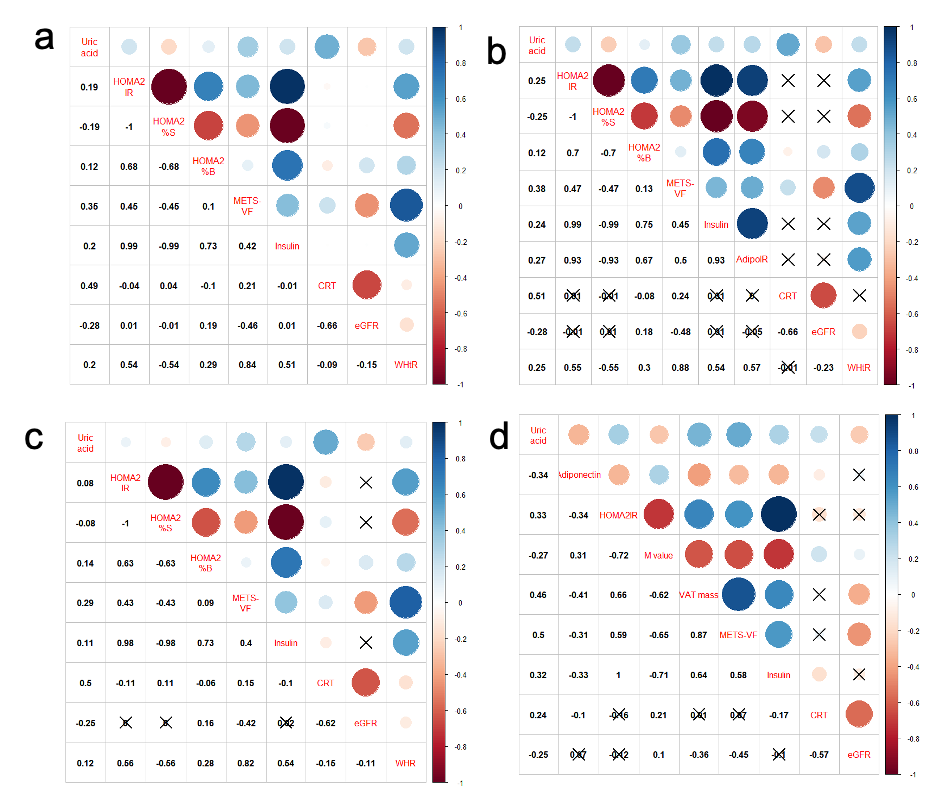

**Supplementary Figure 4:** Correlation matrices for different cohorts. (a) NHANES-ENSANUT joint cohort. (b) NHANES cohort. (c) ENSANUT cohort. (d) UIEM cohort. Abbreviations: AdipoIR: Adipose Insulin Resistance index. CRT: Serum creatinine. eGFR: Estimated glomerular filtration rate. HOMA2-IR:Homeostatic Model for Insulin Resistance. HOMA2%S: Homeostatic Model for Insulin Resistance for pancreatic β cell sensitivity. HOMA2%B: Homeostatic Model for Insulin Resistance for functionality of pancreatic β cells. METS-VF: Metabolic Score for Visceral Fat. WHtR: Weight-height ratio.

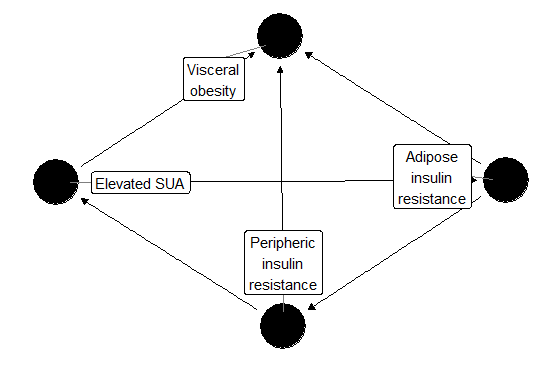

**Supplementary Figure 5:** Directed Acyclic Graph which illustrates the strongest direction of causality for the mechanism.
